# Knowledge Attitude and Practice (KAP of Medications Handling and Storage among Pilgrims during Hajj 2024

**DOI:** 10.1101/2025.06.27.25330399

**Authors:** Bayan Hashim Alsharif, Ghadi Hussain Abdulmajeed, Mohammed Maher Aldurdunji, Mutep Hamed Helal Aljahdal, Sajeed Nazirudheen, Malak Abdulaziz Khiro, Ruaa Hussain Abdulmajeed, Mohammad Kamel AlHarazi, Alhanouf Khalid Alsharif, Khalid Mohammed Alkhalifah, Fai Abdullah Almalki, Thekra Fawaz Albeshri, Khalid Ali Alzahrani, Salma Yonus Alkhayrallah, Saad M. Wali

## Abstract

**Background:** Hajj is the fifth pillar of Islam and one of the largest mass gatherings. The pilgrims with different medical problems may come for hajj with their own medications. Many variables make it difficult to handle and store the medications properly. It may cause degradation of drugs especially the temperature-sensitive medications. Therefore, it is necessary to evaluate the attitude knowledge and practices of Hajj pilgrims regarding medication storage and handling for ensuring the quality, safety, and effectiveness of medications and avoid major risk to one’s health and safety.

**Objective:** to evaluate the knowledge practices and attitude of Hajj pilgrims regarding medications storage and handling during the Hajj season.

**Methods:** The study used cross-sectional design .A survey conducted with electronic questionnaire for data collection by using face to face interview technique. 305 Hajj pilgrims from different nationalities were selected using convenient sampling method. The study was conducted in 2024 among hajj pilgrims.

**Results:** 55.1% of the participants have diabetic mellitus and among them 41.1% used anti-diabetic injectable and 68.1% reported having additional chronic conditions. Knowledge levels indicated that 69.2% of participants had high knowledge scores. Knowledge levels were significantly associated with age, nationality, diabetes status, and use of anti-diabetic injectable. Attitudes toward medication storage practices showed that 80.3% demonstrated positive attitudes. Attitude showed no statistically significant associations with any of the examined variables. Regarding practice levels 52.8% of participants classified as having good practice. Better storage practices were significantly associated with nationality, educational level, and diabetes status.

**Conclusion:** Targeted public education and awareness programs on medication handling among pilgrims are recommended, as medication handling and storage is a sensitive issue and of great importance in daily life.

## Introduction

The fifth pillar of Islam is pilgrimage, or hajj in Arabic (1). One of the biggest mass gatherings occurs when millions of pilgrims from more than 180 nations perform the annual Hajj (2). The holy Kaaba, located in the holy city of Makkah, as well as Mena and Arafat, which are roughly 5 and 18 kilometers away from Makkah, are the three main locations where the Hajj must be performed. It starts on the 8th day of Dhul-Hijjah, the 12th month of the lunar Islamic year, and ends on the 13th day of the same month (3). Travel and movement implied by the Hajj (4). Travel to and from Saudi Arabia for the Hajj takes place via automobiles, buses, trains, and airplanes. Long transit and waiting periods, as well as prolonged flight durations, are all possible. Many outdoor physical activities, sometimes in extremely hot weather, and walking between Makkah and the holy sites of Mina and Arafat are part of the Hajj ceremonies (5). The pilgrimage held in Makkah, Saudi Arabia, which has a scorching desert environment with daily highs of over 25 °C and highs of up to 50 °C (5). Relative humidity ranges from 25% to 50% (4), especially during the summer (5). A considerable portion of the Hajj population is elderly (about 25% are over 65), and many of them have underlying medical issues (6).

It is anticipate that these pilgrims will bring these medications with them on the journey. In case they become necessary, pilgrims also carry additional prescription drugs, such as antibiotics, analgesics, and antipyretics. In the Kingdom of Saudi Arabia (KSA), pilgrims can also obtain drugs from doctors and pharmacies. Through their Hajj medical missions and from other pilgrims they can also obtain medication (5). Many variables make it difficult to handle and store drugs properly during the Hajj, especially for pilgrims who need temperature-sensitive medications that must be stored at cool or low temperatures or for those with chronic diseases who must take their medication on a regular basis (5).

Proper storage conditions are necessary for ensuring the quality, safety, and effectiveness of medicinal items (5). The drugs’ degradation will be accelerated by light, oxygen, humidity, inappropriate temperature and poor storage conditions (7,8). Chemical reactions (such as oxidation from oxygen exposure, hydrolysis from water exposure) and physical responses (such as particle size alteration, suspension/emulsion disintegration, and water adsorption) are the main causes of drug degradation (7). Medication improperly handling, storage, and disposal provide a major risk to one’s health and safety. For patients and those in close proximity, this may occur through the use of pharmaceuticals that are less effective, tainted, or toxic, or through accidental poisoning, as well as for the general public due to environmental contamination caused by pharmaceutical substances that are active (5).

Manufacturers are required to guarantee a potency of 90% to 110% for most medications. The results of stability tests conducted over a specific period of time at a variety of temperatures are used to determine the drug’s expiration date and storage requirements. If a medication is taken by its expiration date and is stored according to suggested guidelines, the manufacturer assures the stability and activity of the product (7). The product packaging insert or the medication label itself contain information about how to store medications. Patients should follow these storage requirements to prevent unfavorable outcomes, including poisoning, exacerbation of underlying health conditions and death, Loss of efficacy of medications can also result in waste and financial loss (5).

With an estimated global prevalence of 9.3% in 2019, diabetes is a significant global public health issue, responsible for sizeable mortality and morbidity worldwide and causing a substantial economic loss. Although many diabetic patients use insulin therapy to manage their condition, lack of knowledge of the therapy and appropriate use of insulin is common and can lead to poor outcomes (12).

The ages, nationalities, cultures, and educational attainment of pilgrims vary, is necessary to evaluate the awareness, knowledge and practices of Hajj pilgrims regarding medication storage and handling during the Hajj mass gathering.

## Materials and methods

This study utilized a cross-sectional survey design to assess the knowledge, attitudes, and practices (KAP) of medication storage among Hajj pilgrims. Ethical approval was obtained from the Institutional Review Board (IRB) of King Abdullah Medical City (KAMC) under approval number 24-1272. The study aimed to evaluate factors influencing medication storage behaviors and identify potential gaps in awareness and adherence to recommended storage practices.

Pilgrims were prospectively recruited for this study through face-to-face interviews. The recruitment period began on 7 June 2024 and ended on 6 July 2024, corresponding to the Hajj season of 1445 AH in the Islamic calendar. Participants approached in person, and only those willing to participate were interviewed. As the study involved the collection of anonymous data through interviews, a verbal informed consent was obtained from each participant prior to the interview. The consent process was conducted privately, with no one else present to ensure confidentiality and voluntariness. The verbal consent was not documented in medical records, as the study did not involve clinical interventions or identifiable health information. No identifiable information was collected from participants. All data were anonymized using unique participant codes (e.g., 001, 002), and participants’ names or other identifiers were neither recorded nor linked to the data. All collected information was securely stored in password-protected electronic files accessible only to the research team.

The survey was pre-tested for clarity and reliability before administration. The survey divided into four key sections. The Demographics and Medical History section included questions about age, gender, nationality, education level, medical expertise, previous Hajj experience, and diabetes status. The Knowledge of Medication Storage section assessed participants’ awareness of proper storage conditions, changes in expiration dates due to improper storage, and the potential health risks associated with incorrect medication handling. The Attitudes Toward Medication Storage section measured participants’ perceptions of the importance of proper medication storage, their willingness to follow recommended guidelines, and their views on the benefits of educational sessions on medication storage. The Medication Storage Practices section examined participants’ adherence to recommended storage conditions, methods of transporting medications, and handling practices during Hajj to ensure medication safety and efficacy.

Scoring methods were applied to assess attitudes, knowledge, and storage practices related to medication management during Hajj. Positive attitudes were measured using four Likert-scale items regarding the importance of checking medications, willingness to follow storage guidelines, perceived benefits of educational sessions, and interest in additional resources, with responses ranging from 1 (Strongly Disagree) to 5 (Strongly Agree), where higher scores indicated more positive attitudes. Knowledge scores were calculated based on three dichotomous questions assessing awareness of medication potency, expiration date changes, and health risks associated with improper storage. Correct answers were assigned a score of 1, while incorrect or uncertain responses received a score of 0, yielding a total score range of 0 to 3, with higher scores reflecting greater knowledge. Storage practice scores were based on two dichotomous questions regarding adherence to proper storage conditions and avoidance of improperly stored medications, with responses coded as 1 for correct practices and 0 for incorrect or uncertain responses, resulting in a total score range of 0 to 2, where higher scores indicated better medication storage practices. Descriptive statistics used to summarize categorical variables as frequencies and percentages, while continuous variables presented as median and interquartile range (IQR). Associations between categorical variables and outcomes assessed using the Chi-square or Fisher’s exact test, as appropriate. Comparisons of continuous variables across groups conducted using the Wilcoxon rank sum test or Kruskal-Wallis test. Multivariable linear regression analyses performed to identify independent predictors of attitudes, knowledge, and storage practices, with results reported as beta coefficients and 95% confidence intervals (CI). A p-value of <0.05 was considered statistically significant. Statistical analyses conducted using RStudio (version 2024.9.1.394, Boston, MA, USA) with R version 4.4.2.

## Results

The study included 305 participants, with the majority aged 45 to <60 years (39.0%), followed by those aged 30 to <45 years (29.8%). More than half of the participants were female (59.0%) and non-Saudi (66.2%). Educational levels varied, with 38.4% holding a bachelor’s degree, 19.7% having completed high school, and 33.1% having a middle school education. Most participants had no medical expertise (83.6%), while 9.8% had basic first aid knowledge, and 6.6% were medical professionals. A quarter of the participants had previously performed Hajj (24.6%). More than half of the participants had been diagnosed with diabetes mellitus (55.1%), and among them, 41.1% used anti-diabetic injectables for diabetes management (**Table 1**).

**Table 1:** Demographic and diabetes-related characteristics.

| Characteristic<sup>1</sup> | Description<sup>2</sup> |
| --- | --- |
| Age (year) |  |
| 18 to <30 | 18 (5.9%) |
| 30 to <45 | 91 (29.8%) |
| 45 to <60 | 119 (39.0%) |
| 60 or more | 77 (25.2%) |
| Gender |  |
| Male | 125 (41.0%) |
| Female | 180 (59.0%) |
| Nationality |  |
| Saudi | 103 (33.8%) |
| Non-Saudi | 202 (66.2%) |
| Educational Level |  |
| Middle school | 101 (33.1%) |
| High school | 60 (19.7%) |
| Bachelor | 117 (38.4%) |
| Master | 19 (6.2%) |
| PhD | 8 (2.6%) |
| Medical Expertise |  |
| None | 255 (83.6%) |
| Basic first aid | 30 (9.8%) |
| Medical professional (e.g., nurse, doctor, etc.) | 20 (6.6%) |
| Previous Hajj Experience | 75 (24.6%) |
| Have been diagnosed with diabetes mellitus | 168 (55.1%) |
| Use of any anti-diabetic injectables to manage diabetes* | 69 (41.1%) |

<sup>1</sup>\*Descriptive data are based on a subset of patients who have been diagnosed with diabetes mellitus (n=168)
<sup>2</sup>n (%)

Among those using anti-diabetic injectables, 65.7% had received education on injectable storage. The majority (76.8%) recognized that storage temperature affects these medications, and 92.3% identified that unused injectables should be stored in a refrigerator. However, only 4.3% knew that used injectables could remain at room temperature for up to four weeks, while 58.0% believed they could be left out for only one day. Almost half (47.8%) were unaware of the signs indicating that an injectable had gone bad, while 24.6% mentioned color change, and 23.2% noted cloudiness as an indicator. Regarding transport to Saudi Arabia, 69.6% carried their injectables in a cooling case, while 23.2% transported them without cooling. Physicians were the primary source of storage instructions (66.7%), followed by pharmacists (15.9%). Most participants (87.0%) knew that insulin should be stored in a refrigerator, and 60.9% correctly stated that insulin could left at room temperature for one day before discarding. When asked how to respond to a refrigerator malfunction during Hajj, 43.5% preferred using a cooling case or seeking medical assistance, while 23.2% would leave the insulin at room temperature, and 24.6% would discard it immediately (**Table 2**).

**Table 2:**
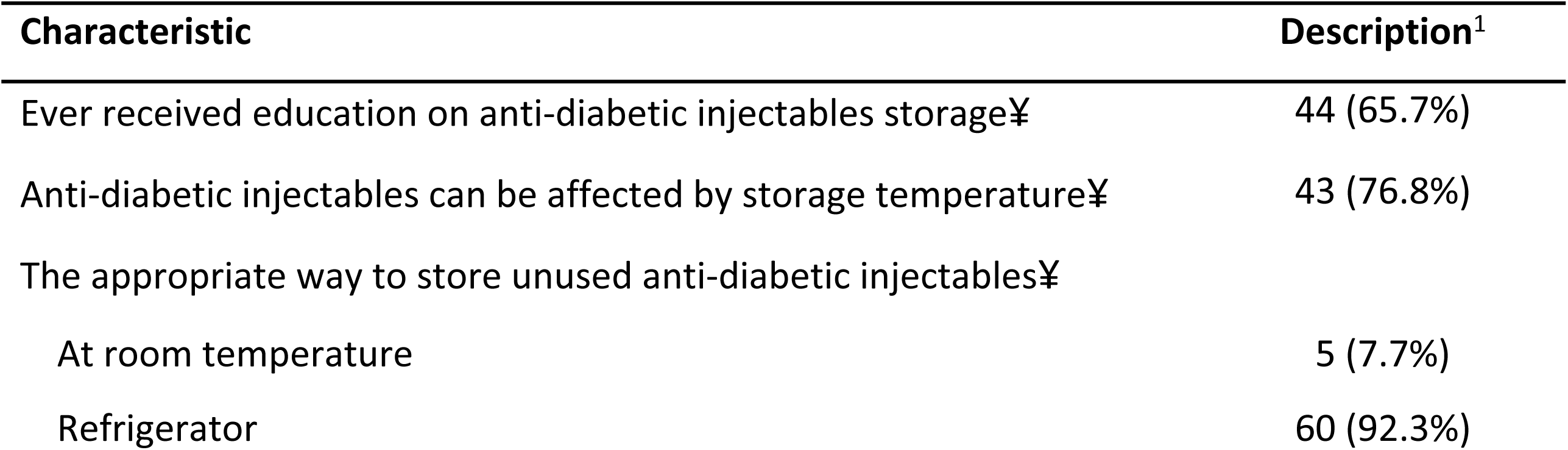
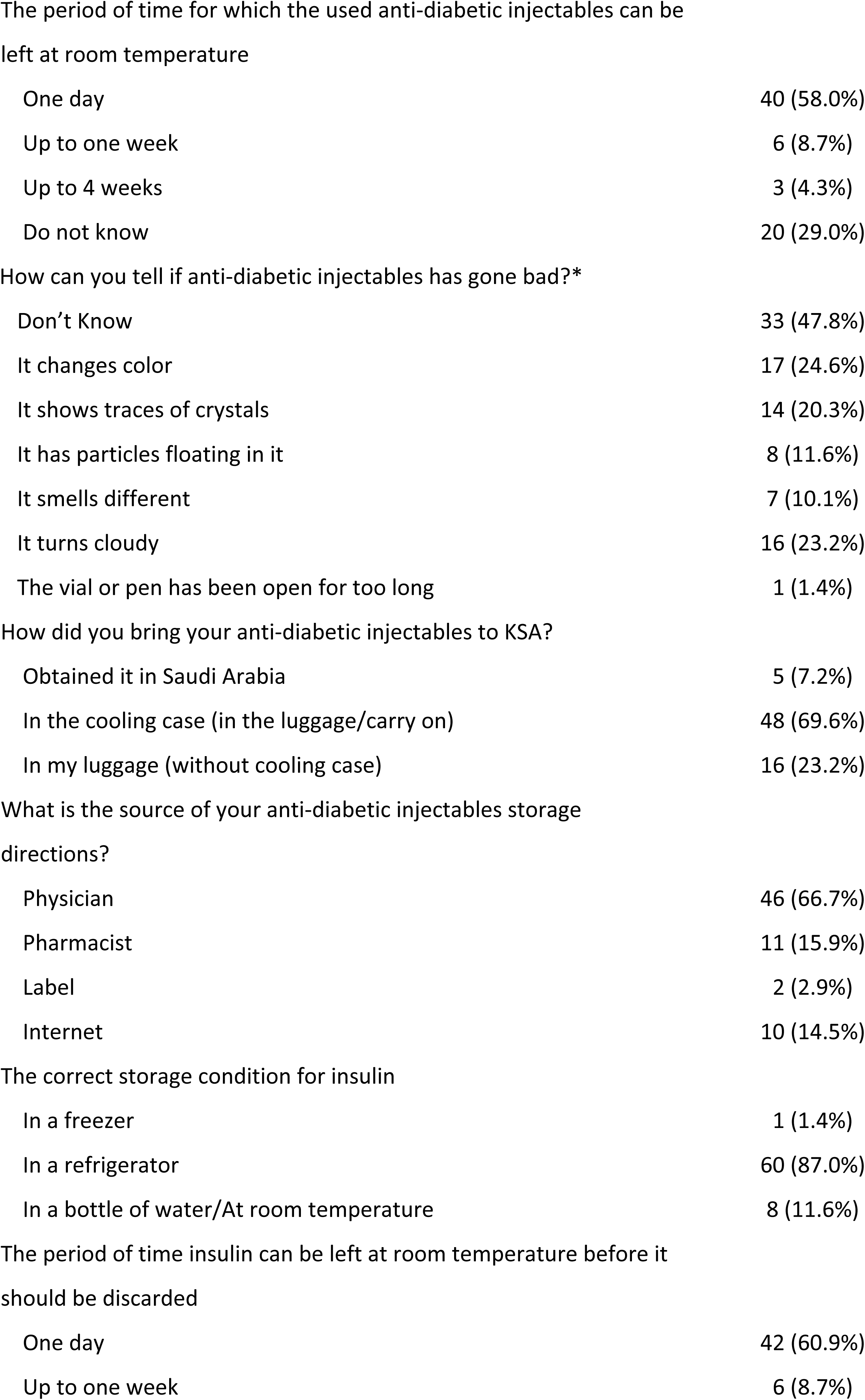
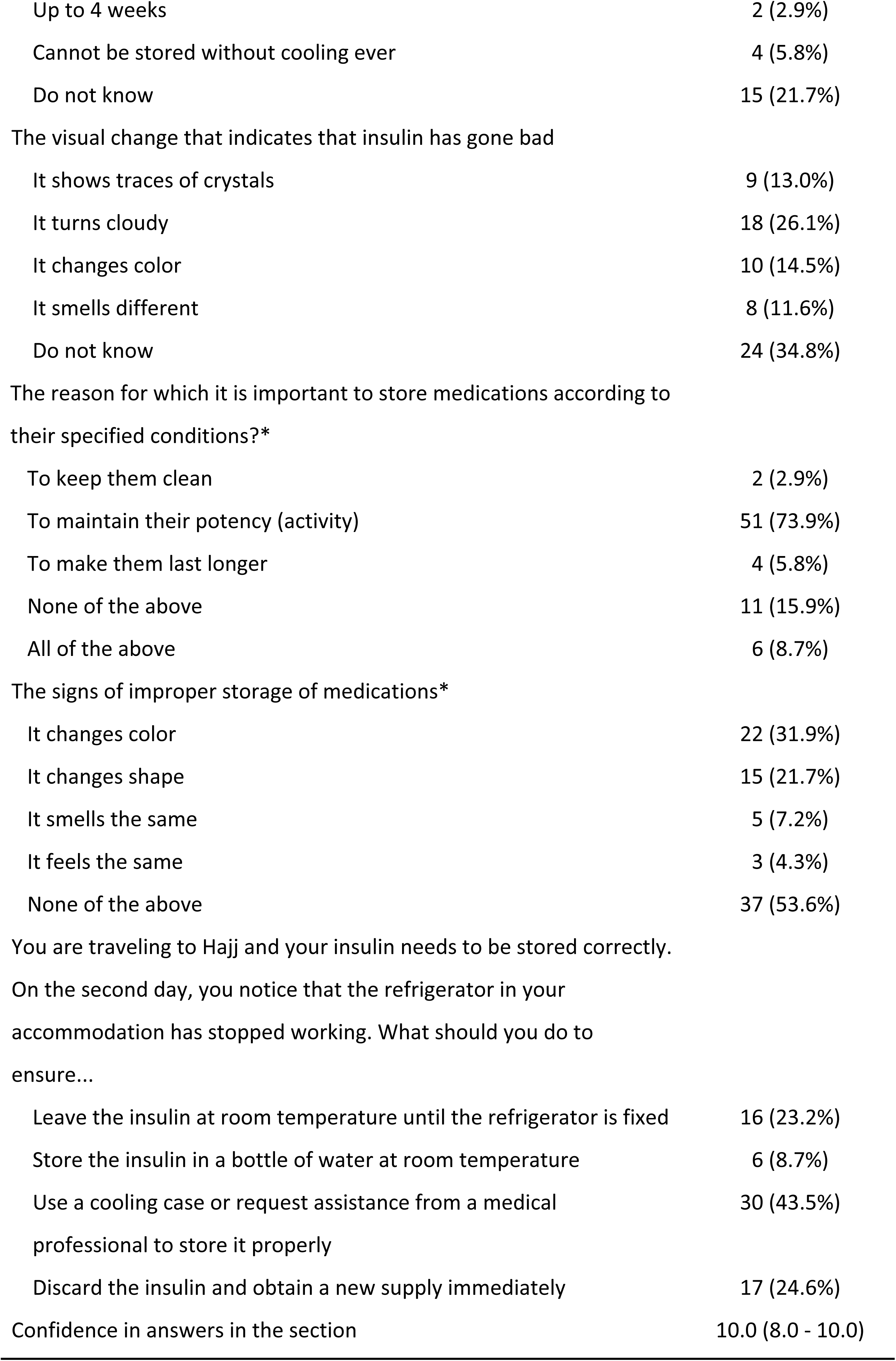
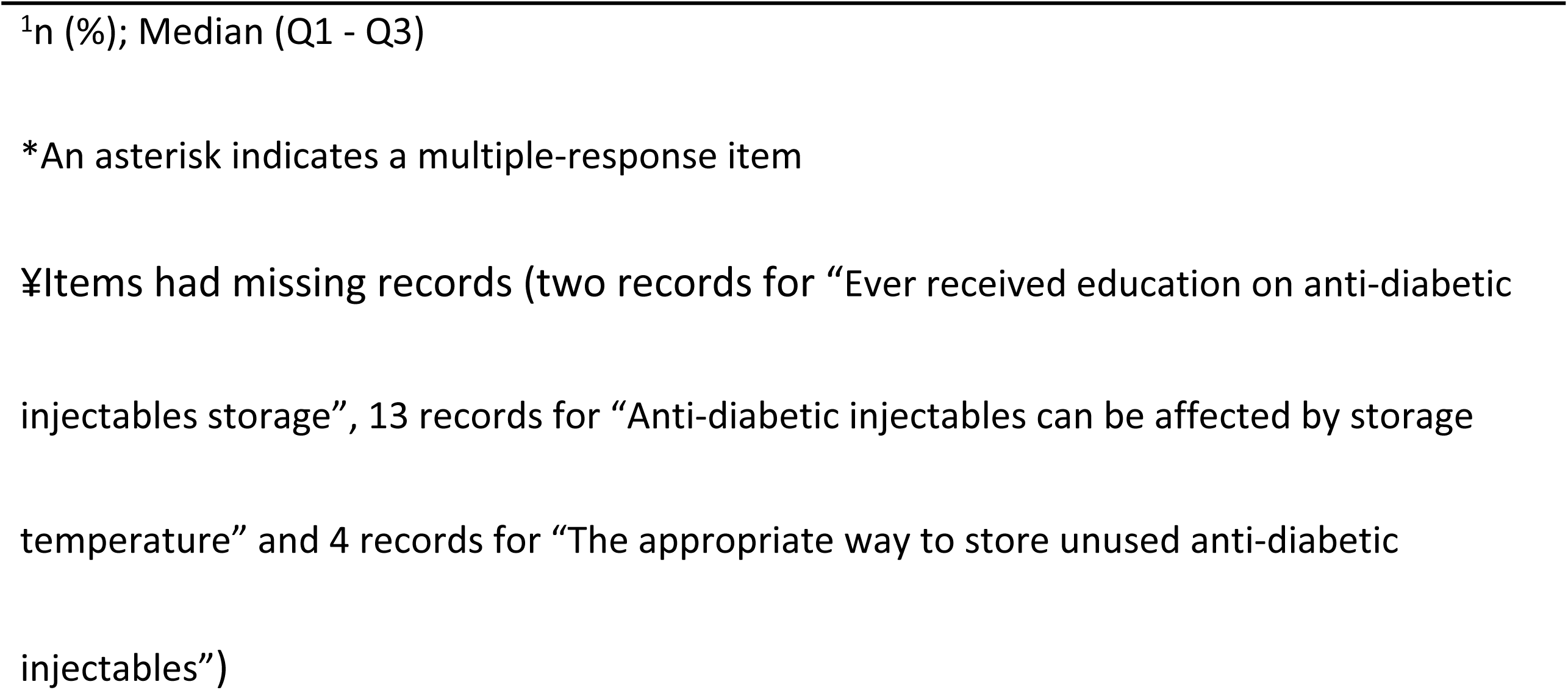
Characteristics of patients who receive anti-diabetic injectables to manage diabetes.

Among individuals with diabetes, 68.1% reported having additional chronic conditions, with hypertension being the most common (70.2%), followed by liver disease (19.1%) and cardiac disease (10.6%). Most participants planned to use one to three medications during Hajj (78.7%), with 72.3% bringing their medications from their home country. During Hajj, 91.5% handled their own medications, and the majority stored them with themselves (68.1%), while 17.0% used a refrigerator, and 10.6% kept medications at room temperature (**Table 3**).

**Table 3:** Characteristics of other health conditions in addition to diabetes mellitus.

| Characteristic | Description<sup>1</sup> |
| --- | --- |
| Have other health conditions in addition to diabetes mellitus | 47 (68.1%) |
| List the chronic diseases* |  |
| Hypertension | 33 (70.2%) |
| Liver disease | 9 (19.1%) |
| Cardiac disease | 5 (10.6%) |
| Kidney disease | 4 (8.5%) |
| Lung disease | 3 (6.4%) |
| Hypercholesterolemia | 2 (4.3%) |
| Asthma | 1 (2.1%) |
| Immunosuppressive conditions | 1 (2.1%) |
| Paracetamol | 1 (2.1%) |
| Hypothyroidism | 1 (2.1%) |
| Rheumatoid arthritis | 1 (2.1%) |
| The number of medications that will be used during Hajj |  |
| 1 to 3 | 37 (78.7%) |
| 4 to 6 | 9 (19.1%) |
| 7 to 9 | 0 (0.0%) |
| 10 or more | 1 (2.1%) |
| The place where you acquire the medications |  |
| Brought from home country | 34 (72.3%) |
| From a healthcare provider or medical mission in KSA | 6 (12.8%) |
| Purchased from a local pharmacy in KSA | 1 (2.1%) |
| Purchased from a hospital or clinic in KSA | 6 (12.8%) |
| Who handles your medication during Hajj? |  |
| Myself | 43 (91.5%) |
| Relative(s) | 4 (8.5%) |
| Where are you going to store you medication during Hajj? |  |
| With myself | 32 (68.1%) |
| At room temperature | 5 (10.6%) |
| In a cooling case | 2 (4.3%) |
| In refrigerator | 8 (17.0%) |

<sup>1</sup>n (%)
\*An asterisk indicates a multiple-response item

The majority of participants agreed or strongly agreed that it is important to regularly check medication conditions during Hajj (83.9%) and that they are willing to follow healthcare professionals’ storage guidelines (86.3%). Similarly, 81.4% found educational sessions on medication storage beneficial, and 75.7% expressed interest in more information on managing medications during Hajj (**Figure 1**). As shown in **Figure 2A**, the majority of participants (80.3%) demonstrated positive attitudes, while 13.1% were neutral and 6.6% had negative attitudes.

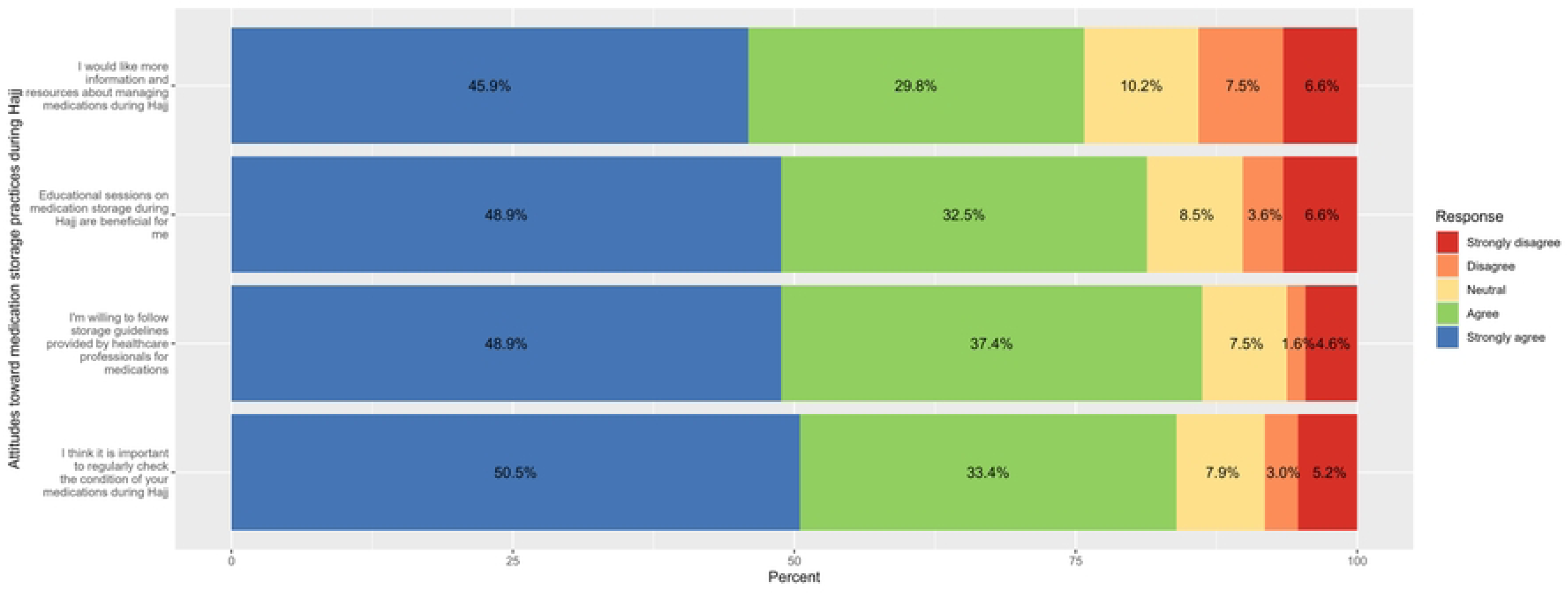

Categorical analysis of attitudes toward medication storage practices showed no statistically significant associations with any of the examined variables. Although not reaching significance, trends were observed in gender and nationality. A higher proportion of males reported positive attitudes (82.4%) compared to females (78.9%) (p = 0.093), and Saudis demonstrated slightly more positive attitudes (83.5%) compared to non-Saudis (78.7%, p = 0.064). Due to the absence of significant predictors, multivariable regression analysis was not performed (**Table 4**).

**Table 4:** Factors associated with attitudes and willingness toward medication storage practices during Hajj.

| Characteristic | Attitudes |  |  | p-value |
| --- | --- | --- | --- | --- |
|  | Negative<br>(Score <10)<br>N=20 | Neutral<br>(Score 10 to 14)<br>N=40 | Positive<br>(Score >14)<br>N=245 |  |
| Age (year) |  |  |  | 0.289 |
| 18 to <30 | 1 (5.6%) | 1 (5.6%) | 16 (88.9%) |  |
| 30 to <45 | 8 (8.8%) | 18 (19.8%) | 65 (71.4%) |  |
| 45 to <60 | 8 (6.7%) | 12 (10.1%) | 99 (83.2%) |  |
| 60 or more | 3 (3.9%) | 9 (11.7%) | 65 (84.4%) |  |
| Gender |  |  |  | 0.093 |
| Male | 11 (8.8%) | 11 (8.8%) | 103 (82.4%) |  |
| Female | 9 (5.0%) | 29 (16.1%) | 142 (78.9%) |  |
| Nationality |  |  |  | 0.064 |
| Saudi | 2 (1.9%) | 15 (14.6%) | 86 (83.5%) |  |
| Non-Saudi | 18 (8.9%) | 25 (12.4%) | 159 (78.7%) |  |
| Educational Level |  |  |  | 0.205 |
| Middle school | 3 (3.0%) | 15 (14.9%) | 83 (82.2%) |  |
| High school | 8 (13.3%) | 11 (18.3%) | 41 (68.3%) |  |
| Bachelor | 8 (6.8%) | 11 (9.4%) | 98 (83.8%) |  |
| Master | 1 (5.3%) | 2 (10.5%) | 16 (84.2%) |  |
| PhD | 0 (0.0%) | 1 (12.5%) | 7 (87.5%) |  |
| Medical Expertise |  |  |  | 0.104 |
| None | 15 (5.9%) | 34 (13.3%) | 206 (80.8%) |  |
| Basic first aid | 4 (13.3%) | 1 (3.3%) | 25 (83.3%) |  |
| Medical professional (e.g., nurse, doctor, etc) | 1 (5.0%) | 5 (25.0%) | 14 (70.0%) |  |
| Previous Hajj Experience | 7 (9.3%) | 9 (12.0%) | 59 (78.7%) | 0.547 |
| Have been diagnosed with diabetes mellitus | 9 (5.4%) | 21 (12.5%) | 138 (82.1%) | 0.582 |
| Use of any anti-diabetic injectables to manage diabetes* | 2 (2.9%) | 12 (17.4%) | 55 (79.7%) | 0.213 |
| n (%) |  |  |  |  |

Fisher's exact test; Pearson's Chi-squared test
A multivariable regression model was not performed due to the lack of significant variables

Results of knowledge levels indicated that 69.2% of participants had high knowledge scores, whereas 30.8% exhibited low knowledge (**Figure 2B**).

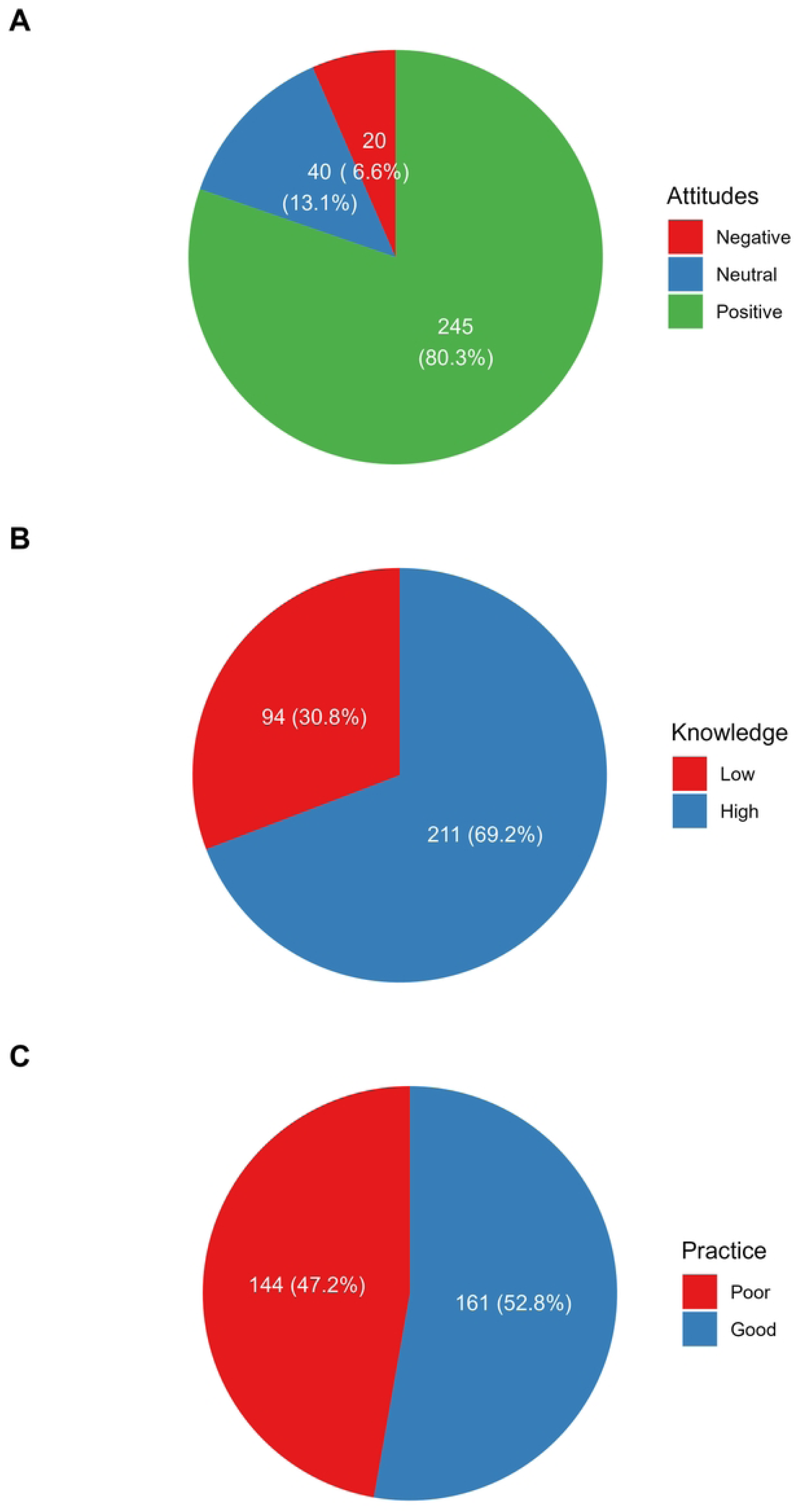

Regarding knowledge, 71.8% correctly identified that medications lose potency if improperly stored, 59.3% recognized that expiration dates could change, and 69.2% understood that inappropriately stored medications could negatively impact health. For storage practices, 61.3% reported storing medications according to specified conditions, while 69.2% admitted to using medications stored inappropriately (**Figure 3**).

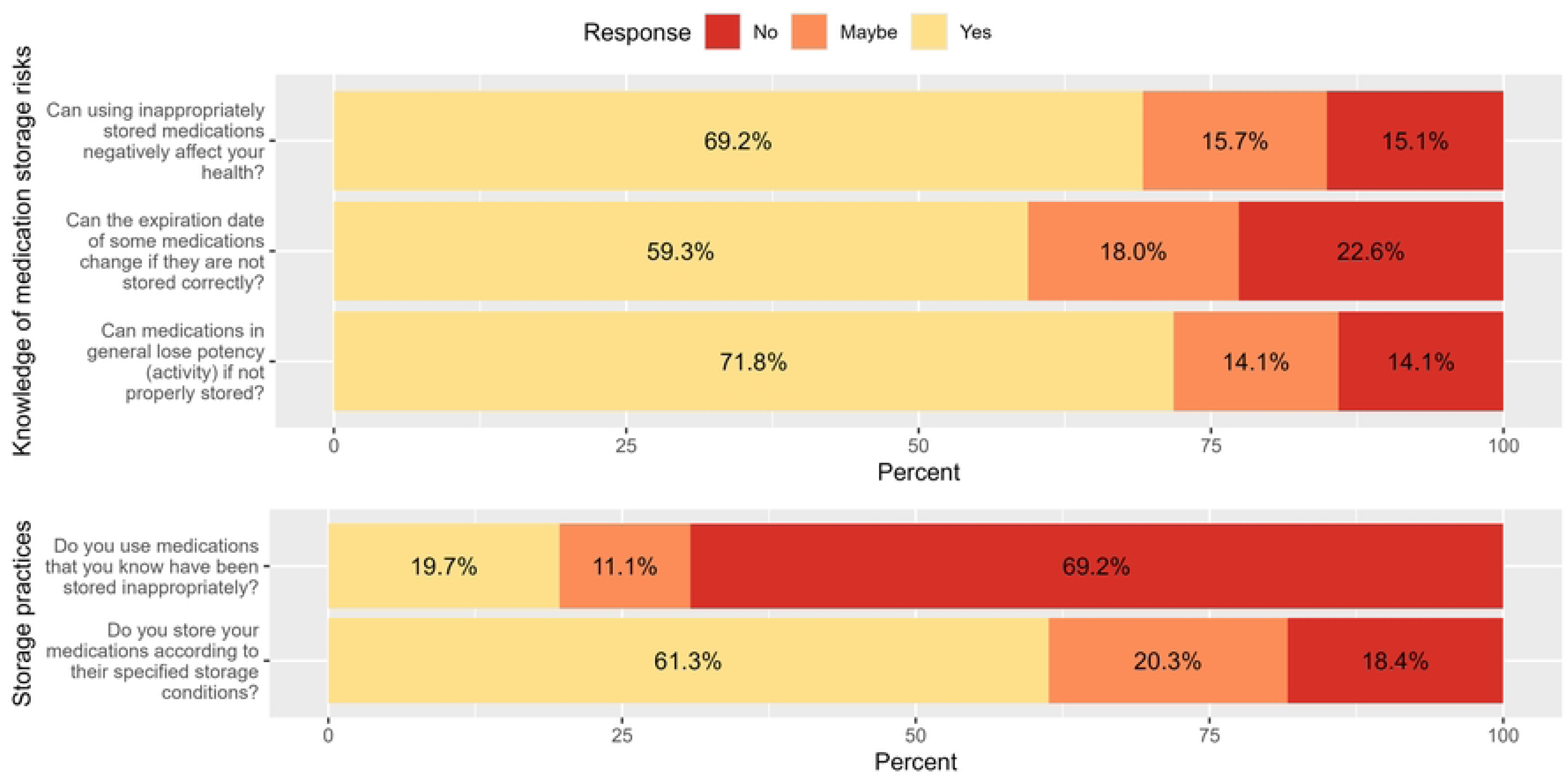

Knowledge levels were significantly associated with age, nationality, diabetes status, and use of anti-diabetic injectables. A higher proportion of participants aged 18 to <30 years had high knowledge levels (94.4%), compared to 55.8% among those aged 60 years or older (p = 0.004). Nationality was a strong predictor, with Saudis more likely to have high knowledge levels (83.5%) compared to non-Saudis (61.9%, p < 0.001); this was confirmed in the multivariable model, where non-Saudi participants had lower odds of high knowledge (OR = 0.31, 95% CI, 0.15 to 0.58, p < 0.001). Diabetes diagnosis was also significantly associated, with non-diabetic participants more likely to have high knowledge (84.7%) than those with diabetes (56.5%, p < 0.001); multivariable analysis confirmed lower odds of high knowledge among diabetics (OR = 0.32, 95% CI, 0.16 to 0.61, p < 0.001). While participants using anti-diabetic injectables had lower knowledge scores (55.1%) compared to those not using them (73.3%, p = 0.004), this association did not remain significant in the multivariable analysis (p = 0.186). Other factors, including gender, educational level, medical expertise, and Hajj experience, were not significantly associated with knowledge scores (**Table 5**).

**Table 5:**
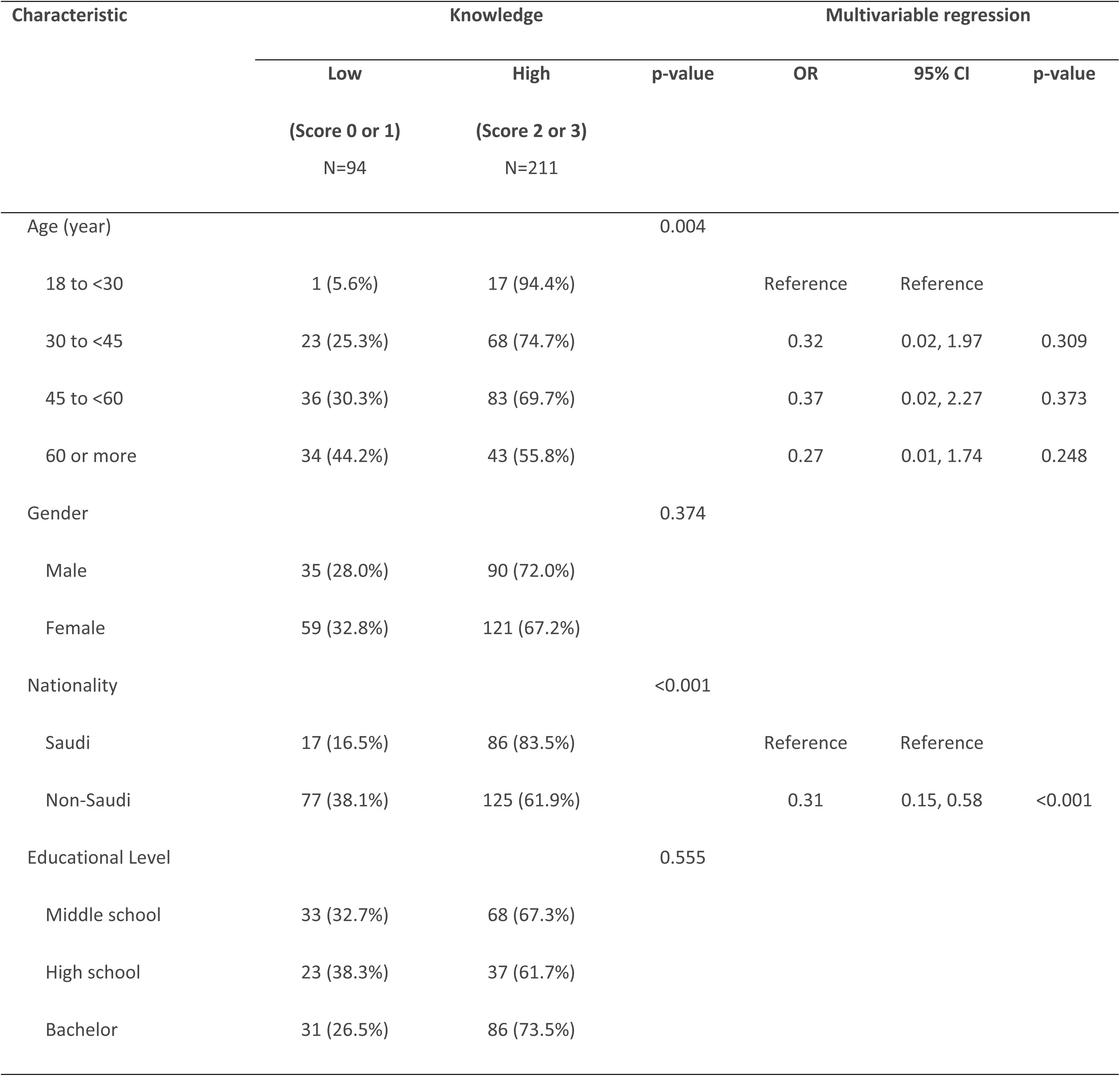
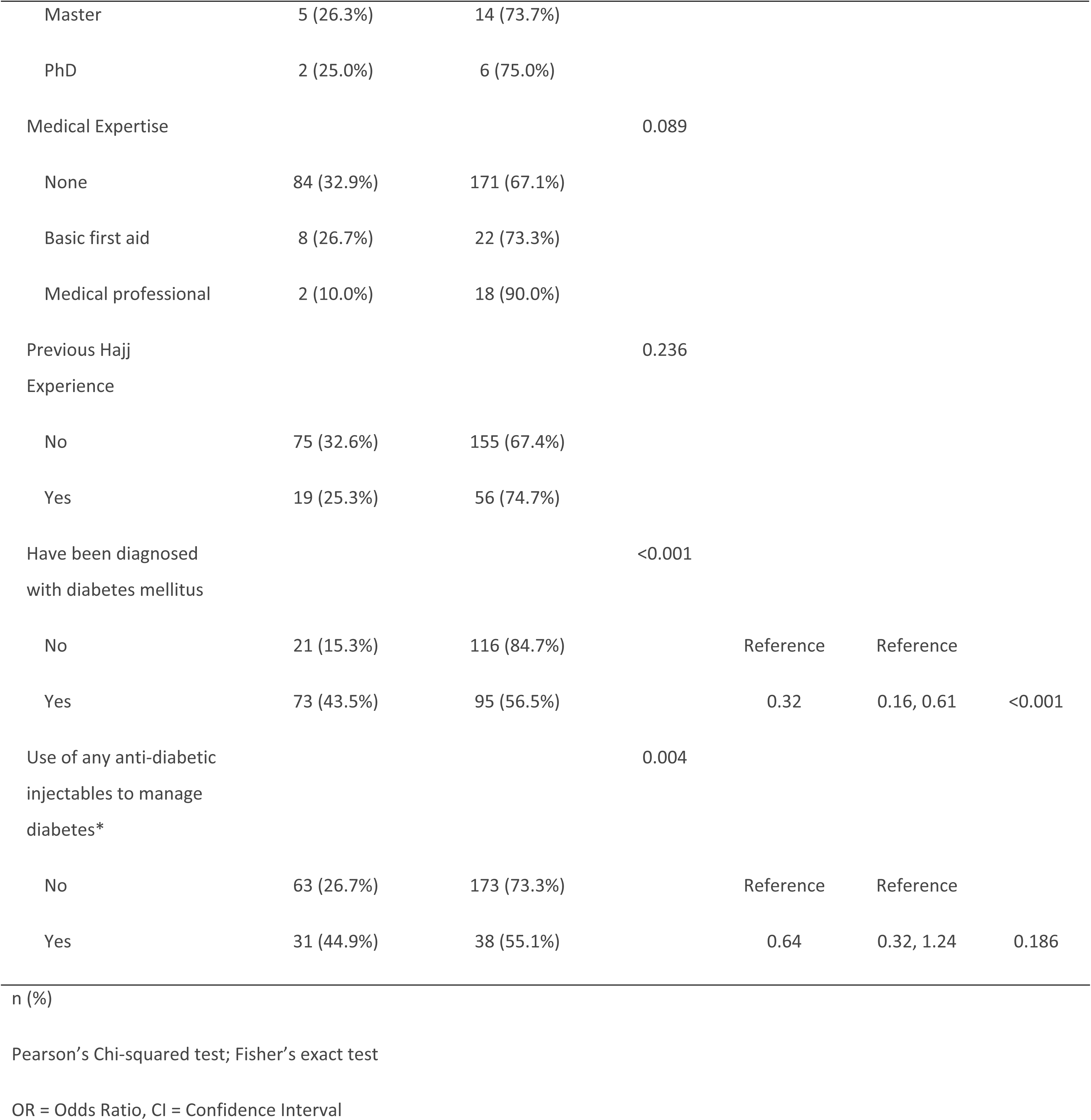
Factors and predictors of higher knowledge scores of medication storage risks.

Regarding practice levels, just over half (52.8%) of participants classified as having good practices and 47.2% as having poor practices (**Figure 2C**).

Better storage practices were significantly associated with nationality, educational level, and diabetes status. A higher proportion of Saudis reported good storage practices (64.1%) compared to non-Saudis (47.0%, p = 0.005), and this association remained significant in the multivariable analysis (OR = 0.54, 95% CI, 0.32 to 0.89, p = 0.016), indicating lower odds of good practices among non-Saudis. Educational level was also significant (p = 0.029); participants with a high school education had the lowest proportion of good practices (35.0%), and were significantly less likely to report good practices compared to those with middle school education (OR = 0.40, 95% CI, 0.20 to 0.78, p = 0.008). Participants diagnosed with diabetes had lower storage practice scores (43.5% reported good practices) compared to those without diabetes (64.2%, p < 0.001), and this association was also significant in multivariable analysis (OR = 0.43, 95% CI, 0.26 to 0.69, p < 0.001). Age, gender, medical expertise, Hajj experience, and use of anti-diabetic injectables were not significantly associated with storage practices (**Table 6**).

**Table 6:** Factors and predictors of having better storage practices.

| Characteristic | Storage practice |  | p-value | Multivariable regression |  |  |
| --- | --- | --- | --- | --- | --- | --- |
|  | Poor | Good |  | OR | 95% CI | p-value |
|  | (Score 0 or 1)<br>N=144 | (Score = 2)<br>N=161 |  |  |  |  |
| Age (year) |  |  | 0.088 |  |  |  |
| 18 to <30 | 7 (38.9%) | 11 (61.1%) |  |  |  |  |
| 30 to <45 | 36 (39.6%) | 55 (60.4%) |  |  |  |  |
| 45 to <60 | 56 (47.1%) | 63 (52.9%) |  |  |  |  |
| 60 or more | 45 (58.4%) | 32 (41.6%) |  |  |  |  |
| Gender |  |  | 0.487 |  |  |  |
| Male | 62 (49.6%) | 63 (50.4%) |  |  |  |  |
| Female | 82 (45.6%) | 98 (54.4%) |  |  |  |  |
| Nationality |  |  | 0.005 |  |  |  |
| Saudi | 37 (35.9%) | 66 (64.1%) |  | Reference | Reference |  |
| Non-Saudi | 107 (53.0%) | 95 (47.0%) |  | 0.54 | 0.32, 0.89 | 0.016 |
| Educational Level |  |  | 0.029 |  |  |  |
| Middle school | 44 (43.6%) | 57 (56.4%) |  | Reference | Reference |  |
| High school | 39 (65.0%) | 21 (35.0%) |  | 0.40 | 0.20, 0.78 | 0.008 |
| Bachelor | 52 (44.4%) | 65 (55.6%) |  | 0.80 | 0.46, 1.40 | 0.443 |
| Master | 6 (31.6%) | 13 (68.4%) |  | 1.49 | 0.52, 4.65 | 0.466 |
| PhD | 3 (37.5%) | 5 (62.5%) |  | 1.34 | 0.30, 6.96 | 0.705 |
| Medical Expertise |  |  | 0.098 |  |  |  |
| None | 126 (49.4%) | 129 (50.6%) |  |  |  |  |
| Basic first aid | 13 (43.3%) | 17 (56.7%) |  |  |  |  |
| Medical professional | 5 (25.0%) | 15 (75.0%) |  |  |  |  |
| Previous Hajj Experience |  |  | 0.240 |  |  |  |
| No | 113 (49.1%) | 117 (50.9%) |  |  |  |  |
| Yes | 31 (41.3%) | 44 (58.7%) |  |  |  |  |
| Have been diagnosed with diabetes mellitus |  |  | <0.001 |  |  |  |
| No | 49 (35.8%) | 88 (64.2%) |  | Reference | Reference |  |
| Yes | 95 (56.5%) | 73 (43.5%) |  | 0.43 | 0.26, 0.69 | <0.001 |
| Use of any anti-diabetic injectables to manage diabetes* |  |  | 0.348 |  |  |  |
| No | 108 (45.8%) | 128 (54.2%) |  |  |  |  |
| Yes | 36 (52.2%) | 33 (47.8%) |  |  |  |  |

Pearson's Chi-squared test; Fisher's exact test
OR = Odds Ratio, CI = Confidence Interval

Participants rated the importance of proper medication storage highly (median = 10.0, IQR = 8.0 to 10.0). Confidence in proper storage and handling was slightly lower (median = 8.0, IQR = 5.0 to 10.0), while concerns about health risks associated with improper storage had a median score of 7.0 (IQR = 2.0 to 10.0). The distributions of scores are depicted in Histograms depicting participants’ responses to items measured on scales from 0 to 10, including the importance of proper medication storage (A), confidence in proper medication storage and handling (B) and concerns about the potential health risks associated with improper medication storage (C) (**Figure 4**)

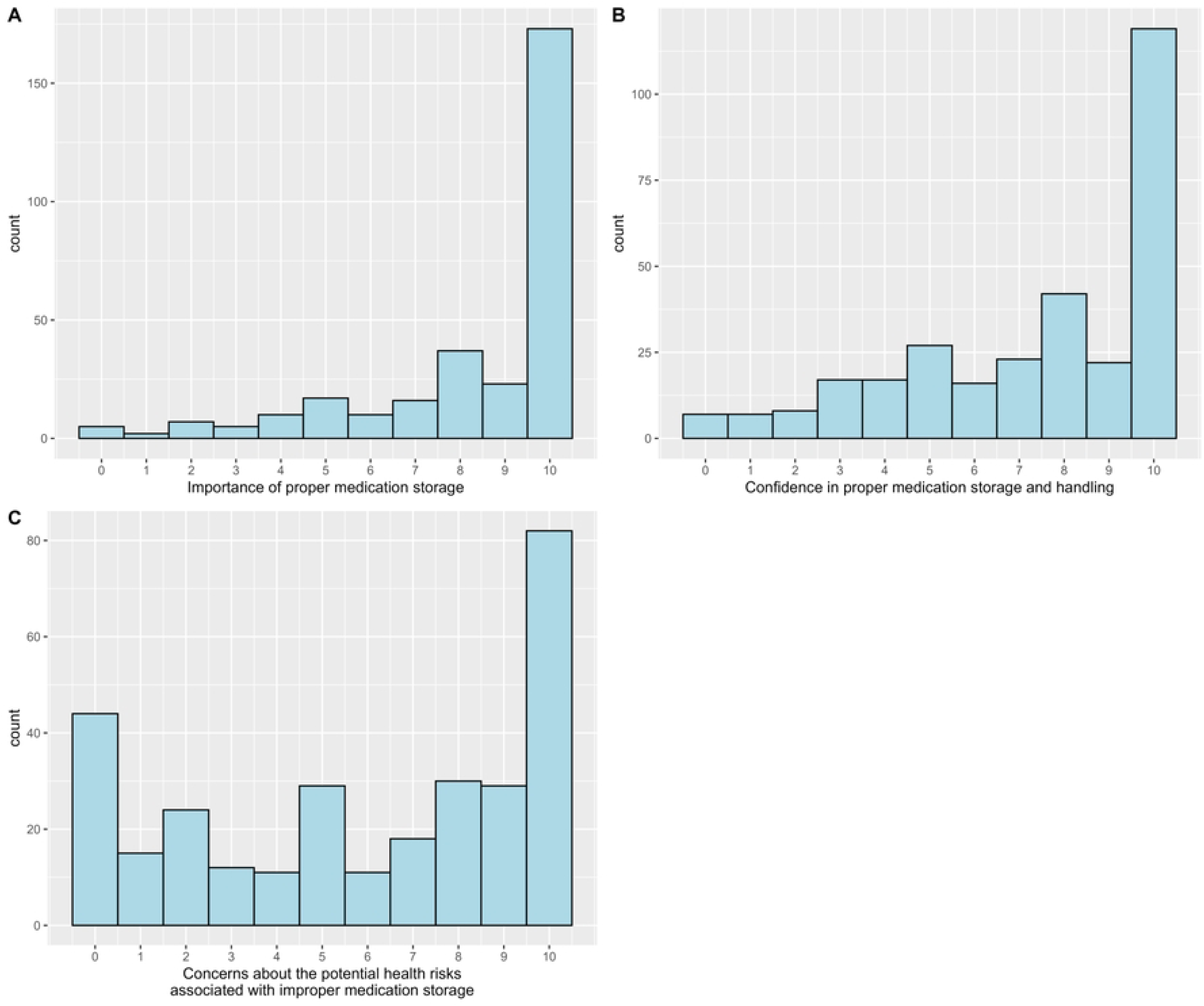

## Discussion

The current study conducted during hajj season of 2023 like many other hajj studies (12). During hajj, the pilgrims are supposed to travel in different vehicles and expose to different environmental changes. In the current study, the participants taken from different area like hospitals, mina, Arafat etc. The temperature of Makkah may be vary between 25 °C to 50 °C (5). These climate changes can cause medication related health impact. (13). Improper storage of some medications like insulin decreases the potency and pharmacological action that medication. Therefore, it is important to have enough knowledge and practice about storage and handling the medications properly (14).

Some objectives of this study found in another studies, but not in detail like current study especially among hajj pilgrims (9) (12). The cross-sectional design is a famous research design and in this study, we use this design for achieving its objectives. The data collected through face-to-face interview (16) with a prepared validated questionnaire like many other studies directed to the pilgrims. Another study use the same design but data collected from the online survey. Both method is reliable and can use for data collection (9)

In the current study, we recruited Hajjis from different places such as hospitals, Makkah ritual site and hotel and Hajj agencies in Makkah after IRB approval. The samples selected by the convenient sampling method. The same sampling method used in many other studies (17). The calculated sample size for this study were 384.but we got only 305 eligible samples during this time. Some studies the participants were more. In the current study, the participants may be less due to limited time as it was hajj study (10). However, another hajj study they took only 277 participants (12). This is as positive factor for this study with comparison of other study sample size.

The survey part of current study had four key sections like (demographic, knowledge, attitude and practice regarding medication storage and handling). Another study with some of similar objectives found with 2 sessions (10) and 3 sessions (12) supporting this study. The four sessions is advantage of this study compare to another studies. SPSS, version 25.0 used for data analysis in this study. The pilot study was conducted clarity and reliability before administration (18)

The ages, nationalities, cultures, and educational attainment of pilgrims vary. It is necessary to evaluate the awareness, knowledge and practices of Hajj pilgrims regarding medication storage and handling during the Hajj mass gathering. In this current study, most of the above-mentioned factors assessed. In current study considerable portion of the participants were elderly like many other hajj studies s (6). Many of the participants have underlying medical issues (6). It may be due to aging and it is a known factor that aging is an inevitable part of life and brings physiologic decline and disease state. (15).In this study majority of the participants were aged between 45 to 60 years (39.0%), but in another study majority were 21-30 years (10). This may be due to large number of elderly population in hajj (6) supported by another study finding (12).

More than half of the participants in this study was females (59.0%) supported by many another studies that the females were 79% (10) and 65.9% (11) respectively. (66.2%) of this study were non-Saudis supported by another hajj study (12). This may due to easy availability of those participants and convenient sampling method. Educational levels shows majority (38.4%) have a bachelor’s degree supported by another study (11). one another study also shows majority were educated (12). This may be due to the Global Educational Reform Movement (19). The study shows only 6.6% were medical professionals. It may be due to global trend in medical profession (20). A quarter of the participants had previously performed Hajj (24.6%). It is personal choice and may vary from time to time.

The manufactures guidelines and drug information assures the stability and activity of the product (7), (5). In this current study only 2.9% of the participants following the labels for the drug information. It may be due to the other factors like the information getting from other sources. The diabetic patient may get the knowledge from different sources regarding the injectable medication (23). The current study shows that main source of information were physicians/nurses about 66.7% and another study have almost similar result (12). This study shows that physicians were the primary source of storage instructions 66.7% followed by pharmacists 15.9%. Another study result shows that physician were the main source of information (12)

Multiple chronic conditions like hypertension, diabetic mellitus, are common among older adults (24). In this study the individuals with diabetes shows that 68.1% of diabetic patient have reported having additional chronic conditions. Among this 70.2%, have HTN followed by liver disease 19.1% and cardiac disease 10.6%. Another study finding says that the prevalence of HTN in adults with diabetes mellitus was 76.3%, 66.0% respectively. The above study definitions and guidelines supporting the current study (25). More than half of the participants has diagnosed with diabetes mellitus and among them, 41.1% used anti-diabetic injectable for diabetes management. Hypertension was the most common comorbidity with diabetes (12). Potentially a large number of pilgrims in each Hajj have diabetes and/or hypertension and other underlying health conditions (6) support this study finding.

Most participants brought one to three medications during Hajj. It may be due taking multiple medications for different disease conditions (24). In this study, 68% of participants keep medications with them supported by another study (10). It is anticipate that these pilgrims may bring some additional medications with them in case they become necessary (5). 78.7% of the participants using 1-3 medications and 19.1% using 4-6 medications. One other study findings shows that pilgrims carry additional medications supporting this study finding (5). Many variables make it difficult to handle and store drugs properly during the Hajj (5). The temperature variance is one of the important concern for storing medications and 76.8% of the current study participants were aware about this.

In this current study, the knowledge levels indicated that 69.2% of participants had high knowledge scores, whereas 30.8% exhibited low knowledge. Some study results shows that the Pilgrims’ knowledge regarding insulin storage was just above average (12) (5). Another one-study result indicated that only 35.6% of participants had good knowledge of insulin pen storage, whereas 64.4% did not (29).

71.8% of the participants in this study correctly identified that medications lose potency if improperly stored, 59.3% recognized that expiration dates could change, and 69.2% understood that inappropriately stored medications could negatively influence health. For storage practices, 61.3% reported storing medications according to specified conditions, while 69.2% admitted to using medications stored inappropriately. Almost similar result found in another study (12). Among those using anti-diabetic injectable, 65.7% had received education on injectable medications and its storage supported by another study (12)

While comparing the knowledge of the participants about injectable diabetic medications the majority 76.8% recognized that storage temperature affects these medications, 92.3% of the participants identified that unused injectable should be stored in a refrigerator. This shows the participants good knowledge about the storage of insulin and it is a positive factor. It was noted that that Pilgrims who were literate and previously received education on insulin storage, those with a higher level of education, and those with a longer duration of insulin therapy, had significantly higher knowledge scores supported by another study (5)

There are different physical signs that indicate signs of bad insulin in the vial (22). While analyzing the current study found that almost half of pilgrims were unaware of the signs indicating that an injectable had gone bad, while 24.6% mentioned color change, and 23.2% noted cloudiness as an indicator. The changes in environment can cause changes damage if the injectable medications. While transportation and movement there is chance of environmental changes. The current study shows that 69.6% carried their injectable in a cooling case and 23.2% transported them without cooling.

Regarding practice levels, just over half (52.8%) of participants classified as having good practices and 47.2% as having poor practices. Another study shows that There was no significant difference in practices between medical professionals and the general population(10). Regarding storage practice scores, nationality, age, medical expertise, and diabetes status were significantly associated with better storage practices. In another study found that four factors that affected the participants’ understanding of insulin-pen storage were age, education level, duration of diagnosis, and years of treatment for diabetes mellitus (21). Proper storage conditions is important factor for ensuring the quality, safety, and effectiveness of medicinal items (5). If not storing properly there is chance for drug degradation (7–8). Improper handling, storage, and disposal of medications is a major risk to one’s health and safety (5). 87% of the participants in this study answered that they will store the insulin in refrigerator.

The majority of participants (80.3%) demonstrated positive attitudes, while 13.1% were neutral and 6.6% had negative attitudes. The majority of participants agreed that it is important to check medication conditions regularly during Hajj and they were willing to follow healthcare professionals’ storage guidelines. This shows that they are following the correct guidelines (26) Another study result shows that The overall patients’ median knowledge and practice levels on insulin storage and handling techniques were moderately adequate and fair respectively (27).

Most of the participants have positive attitude about the insulin storage. 87.0% of this study, participants knew where and how to store insulin. They know importance of education in this area supported by another study (28). Regarding the factors associated with participants’ positive attitudes there noted association with different factors like Saudi nationality, male gender and higher education. The similar studies not found on this area. Similar studies should be conduct to evaluate the attitude in this area.

## Conclusion

In the current study, the hajj pilgrims rated high importance for proper medication storage. However, Confidence in proper storage and handling was slightly lower, while concerns about health risks associated with improper storage had a median score. The current study recommended to conduct targeted public education and awareness programs especially among hajj pilgrims to get more benefit for the benefit of the pilgrims as the medication Handling and Storage is a sensitive issue.

## Data Availability

The data supporting the findings of this study are available from the corresponding author upon reasonable request. Due to ethical restrictions related to participant confidentiality, the full dataset cannot be made publicly available.

## Notes

### Competing Interest Statement

The authors have declared no competing interest.

### Clinical Trial

NA

### Funding Statement

No financial support was received for this study, and the authors have no conflicts of interest to declare.

### Author Declarations

Ethical approval was obtained from the Institutional Review Board (IRB) of King Abdullah Medical City (KAMC) under approval number 24-114 1272.

